# Acute abdominal pain in non-pregnant women presenting to the University Teaching Hospital in Rwanda

**DOI:** 10.1101/2025.09.15.25335784

**Authors:** Turamyimana Faustin, Dushime Jean Paul, Manirafasha Appolinaire, Uwamahoro Doris Lorette

## Abstract

**Introduction:** Acute non-traumatic abdominal pain in non-pregnant women is common in emergency units. Determining its etiology can be challenging, particularly in childbearing age, and requires prompt management. However, women’s abdominal pain has not been studied separately in Rwanda or in the region.

**Objective:** To describe the clinical profile, etiologies, management, and outcomes of non-pregnant women presenting with acute non-traumatic abdominal pain at the Rwanda tertiary hospital.

**Methodology:** Secondary analysis of a prospective cohort at the University Teaching Hospital of Kigali (October 2023 - February 2024). Descriptive statistics, summarized presentation, and etiologies; logistic regression identified predictors for in-hospital mortality.

**Results:** We included 111 women (mean age of 38.3 ±18.1 years; median age 34). At triage, 37.8% were unstable (Orange 33.3%, Red 4.5%). Pain was most often diffuse (53.2%), epigastric (17.1%), hypogastric (10.8%), or right upper quadrant (9.9%). Surgical diagnosis accounted for 46.8% of cases; 27.9%-received interventions, including 26.1% undergoing abdominal surgery. Leading etiologies were intestinal obstruction (20.7%, largely due to adhesions), non-specific abdominal pain (17.1%), peptic ulcer disease (11.7%, gastroenteritis (9.0%), biliary disease or liver abscess (9.0%, peritonitis (8.1%) and malignancy (6.3%). Gynecological causes contributed 8.1% (Adnexal masses 4.5%, Tubo-ovarian abscess 2.7%, and ruptured ectopic pregnancy at 0.9%). Mortality was 8.1% (9/111). Predictors of death included age ≥75 years (OR 7.0; p=0.041, CI: 1.087-45.1), objectified abdominal distension (OR 17.5; p=0.008, CI: 2.1— 145.87), malignancy (OR 16.96; p<0.001, CI: 3.701-77.76), and acute liver injury (OR 6.79; p=0.018 CI: 1.392-33.09).

**Conclusion:** Non-pregnant women with acute abdominal pain frequently present unstable conditions, with nearly half having surgical diagnoses and substantial mortality. Non-specific abdominal pain warrants gynecological assessment and, where available, laparoscopy to avoid missed diagnosis. Broader differential diagnosis, especially in older patients and those with abdominal distension, malignancy, liver injury, and timely surgical or oncological management, may improve outcomes. Strengthening early identification and treatment of high-risk conditions is essential to reducing mortality in this population.

**African Relevance:**

1. Women’s abdominal pain is understudied in Rwanda and wider Africa.
2. Limited diagnostic resources delay recognition of gynecological emergencies.
3. Post-surgical adhesions remain a leading preventable cause regionally.
4. Emergency triage strengthening reduces preventable deaths in African hospitals.
5. Findings inform regional policies on women’s emergency and surgical care.

## Introduction

Acute non-traumatic abdominal pain is of particular concern in emergency medicine, as it is a frequent and complex presenting complaint with a broad differential diagnosis. (1,2) In non-pregnant women, this diagnostic challenge requires careful distinction between gastrointestinal, urological, gynecological, and obstetric etiologies. (3) Gynecological conditions often predominate and include primary dysmenorrhea, endometriosis, mittelschmerz, pelvic inflammatory disease, ovarian cyst accidents, and adhesions. (4)

In Rwanda, acute abdominal pain represents 10.4% of all emergency department (ED) consultations. Although general data are available, the specific etiologies, management approaches, and outcomes for non-pregnant female patients with abdominal pain have not been well characterized in this context. This secondary analysis of a prospective study aims to describe the clinical profile, management, and outcomes of non-pregnant female patients presenting with acute abdominal pain to the ED of a Rwandan tertiary teaching hospital.

## Materials and methods

### Design and Research Setting

The study is a secondary data analysis of a prospective cohort study conducted at the Emergency Department of the University Teaching Hospital of Kigali (CHUK), the largest tertiary hospital in Rwanda. The original cohort, published by Turamyimana et al. in the African Journal of Emergency Medicine, included patients aged 16years or older presenting with acute non-traumatic abdominal pain between October 1, 2023, and February 29, 2024. (5) For this analysis, we excluded male patients and focused exclusively on female patients.

### Study procedure

The original dataset prospectively recorded demographic information, clinical features, investigations, management approaches, complications, and outcomes. For this secondary analysis, data were extracted for females (all presented knowing they are not pregnant) patients. The primary outcome was in-hospital mortality, while secondary outcomes included predictors of death.

### Data analysis

Data were analyzed using the Jamovi platform version 2.7.6.0 (https://www.jamovi.org/). Descriptive statistics were used to summarize baseline characteristics, clinical features, etiologies, management, and outcomes. Binomial logistic regression was used to assess predictors of mortality. A p-value <0.05 was considered statistically significant.

### Ethical Consideration

This secondary analysis used data from a prospective cohort study conducted at the Emergency Department of the University Teaching Hospital of Kigali (CHUK) between October 1, 2023, and February 29, 2024. The original study was approved by the Institutional Review Board of the College of Medicine and Health Sciences, University of Rwanda (Approval No. 370/CMHS IRB/2023), and by the Ethics Committee of CHUK (Approval Ref. EC/CHUK/159/2023). Both committees reviewed and approved the informed consent process. Written informed consent was obtained from all participants, covering study purpose, voluntary participation, confidentiality, and the right to withdraw without affecting their care. This secondary analysis was conducted under the same approvals.

## Result

Among these, 261 patients were included in the original study, and 111 patients were females. The mean age for female patients was 38.3 (±18.1 standard deviation). Triage category using the Rwanda modified South African Triage Scale, locations, symptoms, past history, clinical findings, and etiology are summarized in Table 1.

**Table 1.**
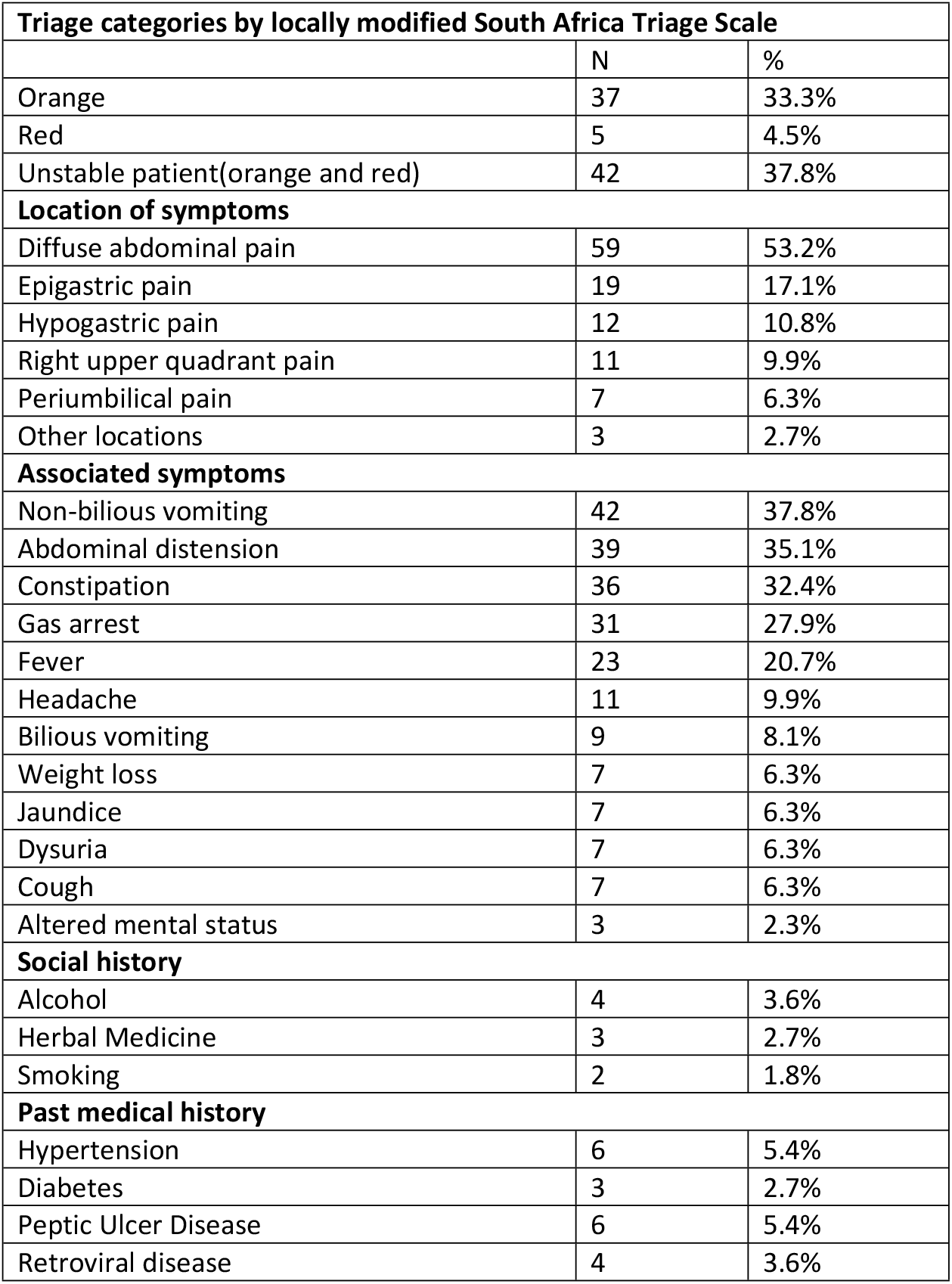
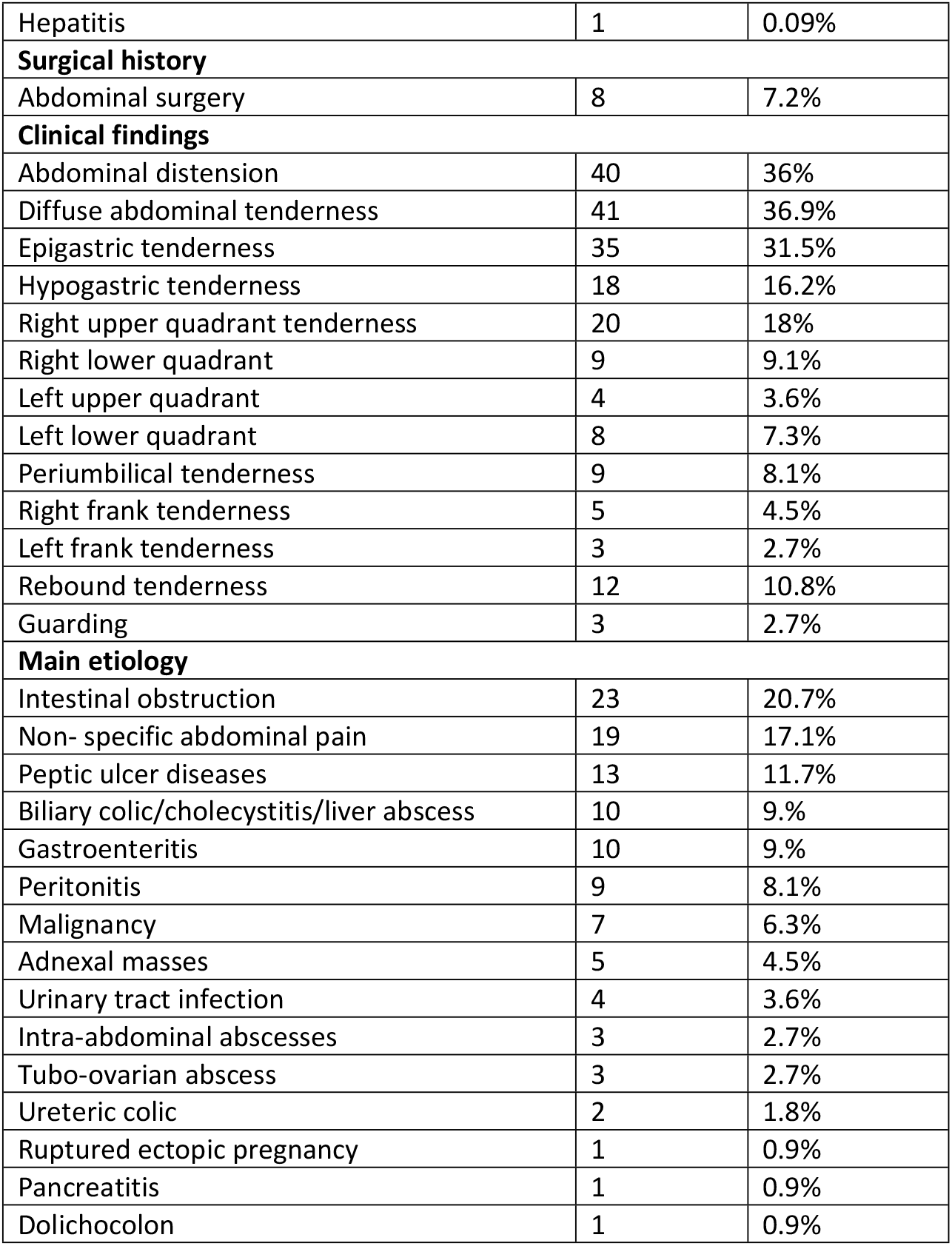
Baseline characteristics and clinical features of non-pregnant women with acute abdominal pain.

Among non-pregnant women with abdominal pain, 46.8% (n=52) had surgical conditions (Table 1). Interventional management was required in 27.9% of patients, including 26.1% (n=29) who underwent abdominal surgery, one who had a colonoscopy, and one who had cystoscopy with bilateral stent placement. Gynecological causes accounted for 8.1% of cases, including adnexal masses (4.5%), tubo-ovarian abscesses (2.7%), and 1 case of ruptured ectopic pregnancy (0.9%). Overall, 18% (n=20) were managed conservatively. Most non-surgical cases were treated as outpatients (30.6%, n=34), while 19.8% (n=22) required admission to internal medicine. A small group (n=5) was managed in the Emergency Department and discharged home, including two cases of intestinal obstruction treated conservatively and three medical cases.

### Outcome measures

Among the 111 patients, nine died, giving a mortality rate of 8.1%. Factors significantly associated with death are summarized in Table 2. Age ≥75 years (OR 7.0; 95% CI 1.09–45.1; p=0.041), abdominal distension (OR 17.5; 95% CI 2.10–145.9; p=0.008), malignancy (OR 16.96; 95% CI 3.70–77.8; p<0.001), and acute liver injury (OR 6.79; 95% CI 1.39–33.1; p=0.018) were independently associated with mortality.

**Table 2.** Risk factors for hospital mortality.

| Risk factor | Odd ratio | 95% Confidence interval | p-value |
| --- | --- | --- | --- |
| Age $\geq 75$ years | 7.0 | 1.087- 45.1 | 0.041 |
| Distended abdomen | 17.5 | 2.10– 145.87 | 0.008 |
| Malignancy | 16.96 | 3.701 – 77.76 | <0.001 |
| Acute liver injury | 6.79 | 1.392 – 33.09 | 0.018 |

## Discussion

Abdominal pain in women is a common presentation among women in the emergency department, with etiologies including both gynecological and non-gynecological conditions(3,6). In our study, non-gynecological predominated, with intestinal obstruction and other gastrointestinal disorders accounting for 60.3% of cases. This contrasts with the findings of Gilling Smith et al, where gynecological conditions were more frequent(3). There are similar findings to our study by Hagipoglu et al., where gastrointestinal conditions were predominate cause in two-third of cases. (6) Nonspecific abdominal pain was less common in other studies; however, it may mask an underlying specific diagnosis, as noted by Gaitan et al. (7) Mortality in our study cohort was high but comparable to the regional rate, such as in Tanzania(8), but mostly related to high age, distended abdomen, malignancy, and acute liver injury. This mortality is higher than that found in Germany, of 2.6% exclusive for female patients. (9)

These findings provide insight into the burden and outcomes of acute abdominal pain among non-pregnant women in Rwanda.

### Limitations of the study

This secondary analysis is subject to several limitations. First, selection bias is possible since the most severely ill patients were often unable to provide consent; inclusion of such patients might have led to even higher mortality estimates. Second, as a single-center study from a tertiary-care teaching hospital, the findings may not be generalizable to rural or lower-level facilities. Third, the analysis was restricted to variables collected in the original study, meaning that comorbidities, socioeconomic status, and healthcare-seeking behavior could not be assessed. Subgroup analyses further reduced the sample size, limiting statistical power. Finally, we did not follow patients after ED discharge, so outcomes such as repeat visits, complications, or deaths in the community were not captured. Despite these limitations, the prospective cohort design, standardized triage, and focus on a previously understudied group of non-pregnant women with acute abdominal pain strengthen the validity and relevance of our findings.

## Conclusion

Abdominal pain complaints in women are a common presentation in Rwandan emergency departments, with nearly half of the patients having had surgical conditions. Only about a quarter required interventional management (27.9%), most of whom underwent abdominal surgery (26.1%). While non-gynecological causes predominated, gynecological conditions still represented a meaningful proportion and should not be overlooked. Patients with non-specific abdominal pain warrant careful evaluation with gynecological consultation or laparoscopic assessment to uncover otherwise missed pathology. Women of reproductive age should always be screened for pregnancy and investigated for tubo-ovarian or adnexal pathology. The predominance of intestinal obstruction, largely due to post-surgical adhesions, underscores the need for preventive strategies and vigilant postoperative follow-up. Early recognition and prioritization of high-risk patients, particularly older women and those presenting with abdominal distension, malignancy, or acute liver injury, may enable more timely surgical and oncologic interventions and help reduce mortality in resource-limited settings.

Although these findings come from a single tertiary center and should be interpreted with caution, given potential selection bias and absence of post-discharge follow-up, the prospective, systematically collected dataset from the country’s largest referral hospital strengthens the validity and regional relevance of the results.

## Dissemination of results

We will share the results of this secondary analysis mainly through academic and professional channels. The findings will appear in peer-reviewed journals and be presented at scientific conferences, both internationally and regionally, to help expand knowledge about abdominal pain in emergency departments. Locally, we will communicate the results to CHUK leadership, the Ministry of Health, and clinical teams to support policy and practice improvements. We will also use the findings in academic discussions and training programs to strengthen emergency care in Rwanda and similar settings.

## Data Availability

All relevant data are within the manuscript and its Supporting Information files.

## CRediT authorship contribution statement

**Faustin Turamyimana:** Conceptualization, Methodology, Project administration, Investigation, Data curation, Formal analysis, Resources, Writing – original draft, Writing – review & editing, Visualization, Funding acquisition. **Jean Paul Dushime:** Validation, review & editing. **Doris Lorette Uwamahoro:** Validation. **Apollinaire Manirafasha**: Validation

## References

1. Gennaro P. “Acute Abdomen in Women of Childbearing Age: Appendicitis or Pelvic Inflammatory Disease? A Systematic Review.” Biomed J Sci Tech Res [Internet]. 2021 May 10;35(4):27900–6. Available from: https://biomedres.us/fulltexts/BJSTR.MS.ID.005738.php

2. Fagerström A, Paajanen P, Saarelainen H, Ahonen-Siirtola M, Ukkonen M, Miettinen P, et al. Non-specific abdominal pain remains the most common reason for acute abdomen: 26-year retrospective audit in one emergency unit. Scand J Gastroenterol [Internet]. 2017 Oct 3;52(10):1072–7. Available from: 10.1080/00365521.2017.1342140

3. Gilling-Smith C, Panay N, Wadsworth J, Beard RW, Touquet R. Management of women presenting to the accident and emergency department with lower abdominal pain. Ann R Coll Surg Engl. 1995 May;77(3):193–7.

4. Jivraj S, Farkas A. Gynaecological causes of abdominal pain. Surg [Internet]. 2015;33(5):226–30. Available from: https://www.sciencedirect.com/science/article/pii/S0263931915000459

5. The jamovi project (2025). [Internet]. Vol. 15, African Journal of Emergency Medicine. 2025. p. 100895. Available from: https://www.sciencedirect.com/science/article/pii/S2211419X25000357

6. Hatipoglu S, Hatipoglu F, Abdullayev R. Acute right lower abdominal pain in women of reproductive age: clinical clues. World J Gastroenterol. 2014 Apr;20(14):4043–9.

7. Gaitán HG, Reveiz L, Farquhar C, Elias VM. Laparoscopy for the management of acute lower abdominal pain in women of childbearing age. Cochrane database Syst Rev. 2014 May;2014(5):CD007683.

8. Mjema KM, Sawe HR, Kulola I, Mohamed AS, Sylvanus E, Mfinanga JA. Aetiologies and outcomes of patients with abdominal pain presenting to an emergency department of a tertiary hospital in Tanzania: a prospective cohort study. BMC Gastroenterol. 2020;20(1):16.

9. Helbig L, Möckel M, Fischer-Rosinsky A, Slagman A. Non-Traumatic Abdominal Pain. Vol. 120, Deutsches Arzteblatt international. Germany; 2023. p. 613–4.

